# Recovered not restored: Long-term health consequences after mild COVID-19 in non-hospitalized patients

**DOI:** 10.1101/2021.03.11.21253207

**Authors:** Max Augustin, Philipp Schommers, Melanie Stecher, Felix Dewald, Lutz Gieselmann, Henning Gruell, Carola Horn, Kanika Vanshylla, Veronica Di Cristanziano, Luise Osebold, Maria Roventa, Toqeer Riaz, Nikolai Tschernoster, Janine Altmueller, Leonard Rose, Susanne Salomon, Vanessa Priesner, Jan Christoffer Luers, Christian Albus, Stephan Rosenkranz, Birgit Gathof, Gerd Fätkenheuer, Michael Hallek, Florian Klein, Isabelle Suárez, Clara Lehmann

## Abstract

**Background:** While the leading symptoms during coronavirus disease 2019 (COVID-19) are acute and the majority of patients fully recover, a significant fraction of patients now increasingly experience long-term health consequences. However, most data available focus on health-related events after severe infection and hospitalization. We present a longitudinal, prospective analysis of health consequences in patients who initially presented with no or minor symptoms of severe acute respiratory syndrome coronavirus type 2 (SARS-CoV-2) infection. Hence we focus on mild COVID-19 in non-hospitalized patients.

**Methods:** We included 958 patients with confirmed SARS-CoV-2 infection in this study. Patients were observed for seven months from April 6^th^ to December 2^nd^ 2020 for long-term symptoms and SARS-CoV-2 antibodies. We identified anosmia, ageusia, fatigue or shortness of breath as most common, persisting symptoms at month 4 and 7 and summarized presence of such long-term health consequences as post-COVID syndrome (PCS). Predictors of long-term symptoms were assessed using an uni- and multivariable logistic regression model.

**Findings:** We observed 442 and 353 patients over four and seven months after symptom onset, respectively. Four months post SARS-CoV-2 infection, 8.6% (38/442) of patients presented with shortness of breath, 12.4% (55/442) with anosmia, 11.1% (49/442) with ageusia and 9.7% (43/442) with fatigue. At least one of these characteristic symptoms was present in 27.8% (123/442) and 34.8% (123/353) at month 4 and 7 post-infection, respectively. This corresponds to 12.8% patients with long-lasting symptoms relative to the initial total cohort (123/958). A lower baseline level of SARS-CoV-2 IgG, anosmia and diarrhea during acute COVID-19 were associated with higher risk to develop long-term symptoms.

**Interpretation:** The on-going presence of either shortness of breath, anosmia, ageusia or fatigue as long-lasting symptoms even in non-hospitalized patients was observed at four and seven months post-infection and summarized as post-COVID syndrome (PCS). The continued assessment of patients with PCS will become a major task to define and mitigate the socioeconomic and medical long-term effects of COVID-19.

**Funding:** COVIM:„NaFoUniMedCovid19”(FKZ: 01KX2021)

**Research in context:** *Evidence before this study:* Data about long-term health consequences after SARS-CoV-2 infection and COVID-19 is scarce and most available data describe health consequences in hospitalized patients during acute COVID-19. However, these studies do not take into account the vast majority of patients with a milder course of infection (WHO score1-3).

*Added value of this study:* Our cohort consists of mostly mild COVID-19 cases that have been prospectively followed for a median time of 6.8 months. At least one trained physician critically reviewed the patients’ reported symptoms at each visit. We assessed SARS-CoV-2 IgG at each visit to correlate reported symptoms with serological data. At 4 months after SARS-CoV-2 infection, shortness of breath occurred in 8.6% (38/442), anosmia in 12.4% (55/442), ageusia in 11.1% (49/442), and fatigue in 9.7% (43/442) of patients. At least one characteristic symptom was present in 27.8% (123/442) and 34.8% (123/353) at months 4 and 7 post-infection, respectively. Symptoms were summarized as post-COVID syndrome (PCS). Relative to our initial total cohort (123/958), this corresponds to 12.8% patients with long-lasting symptoms. Lower baseline level of SARS-CoV-2 IgG, anosmia and diarrhea during acute COVID-19 were associated with higher risk of developing long-term symptoms.

*Implications of all available evidence:* We believe that our findings have important implications for the fields of infectious diseases and public health, because we show long-term health consequences may occur even after very mild COVID-19 in the outpatient setting. As up to 81% of all SARS-CoV-2 infected patients present with mild disease, it can be expected that PCS will affect a larger number of individuals than initially assumed, posing major medical, social and economic challenges.

## Introduction

As the number of severe acute respiratory syndrome coronavirus type 2 (SARS-CoV-2) infections sharply increased during the coronavirus disease 19 (COVID-19) pandemic in March 2020 ^1^, we established a post-COVID outpatient clinic at the University Hospital Cologne (UHC) in Germany. Starting on April 6^th^ 2020, SARS-CoV-2-convalescent patients were invited to present regularly to longitudinally investigate the development of SARS-CoV-2 immunity, screen for potential plasma donation and perform check-up visits on COVID-19. By June 2020, we observed that substantial numbers of SARS-CoV-2-convalescent patients complained of symptoms months after acute infection and had not returned to their initial health state prior to infection, leading us to suspect a possible post-COVID syndrome (PCS).

Data about the longitudinal health-related events is scarce and most available data describe patients that have been hospitalized during acute COVID-19 ^2^. However, with only about 5-10% ^3^ of patients being hospitalized, these studies do not take into account the vast majority of patients with milder course of infection (WHO score 1-3). Hence, we set out to exploring the incidence, diagnostic criteria and management of long-term health consequences at 4 and 7 months after mild courses of COVID-19.

## Methods

### Study design and setting

In this prospective and longitudinal study, we included SARS-CoV-2-convalescent patients, who presented to the post-COVID outpatient clinic of UHC from April 6th onwards (figure 1). To reach as many convalescent plasma donors as possible in a short period of time, attention was drawn to the project through public media. Subsequently, numerous interested individuals contacted us to participate in the study.

**Figure 1.**
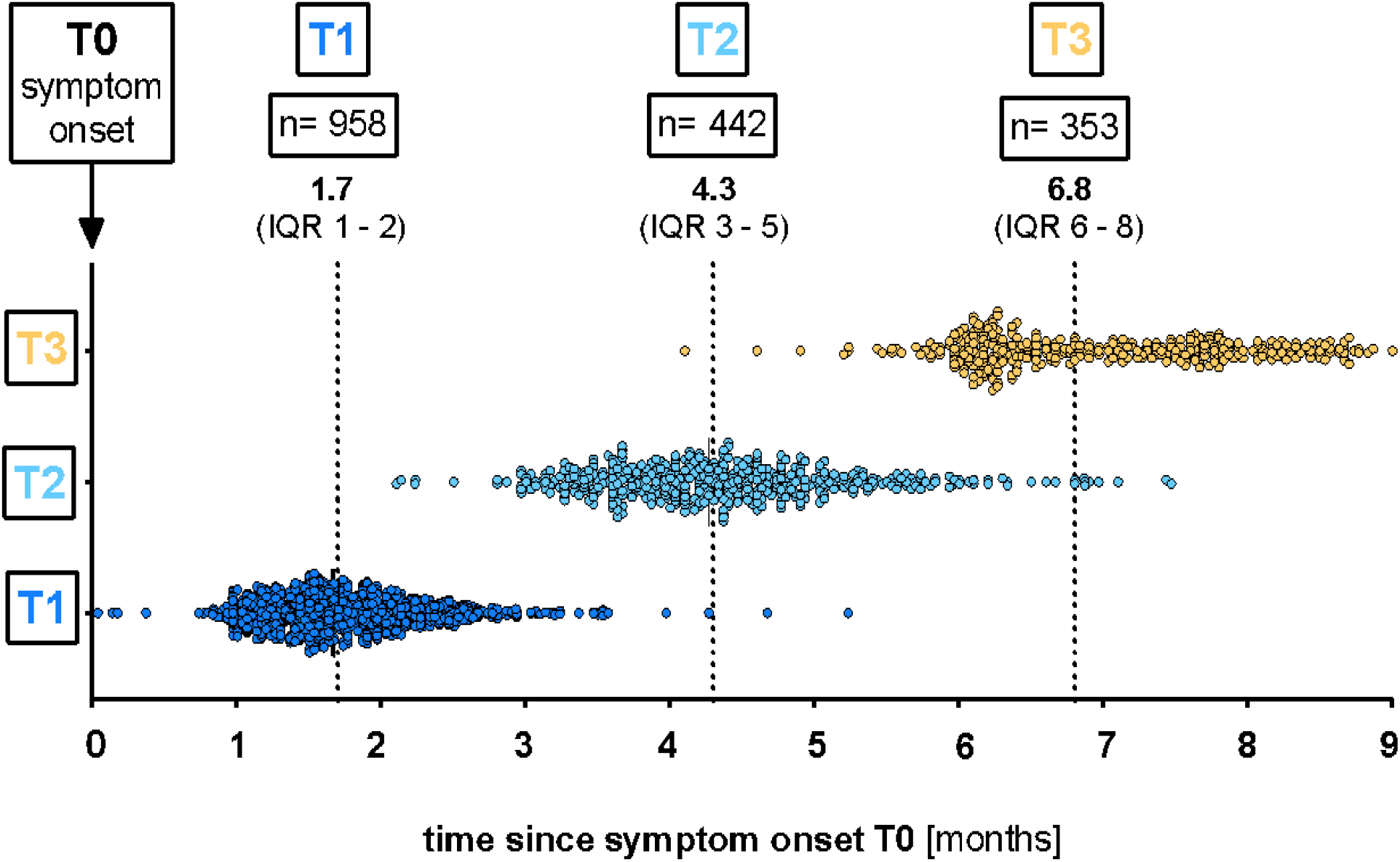
Total visits at the post-COVID outpatient clinic of the UHC April 6^th^ till December 2^nd^ 2020. Median time (months) of the respective visit (T1, T2 and T3) after the onset of symptoms (T0): T1 1.7 (IQR 1.4-2.1), T2 4.3 (IQR 3.7-4.9) and T3 6.8 (IQR 6.2-7.8). UHC, University Hospital Cologne; IQR, interquartile range; T0, symptom onset; T1, first study visit; T2 second study visit; T3, third study visit.

### Participants and eligibility criteria

Inclusion criteria were: (1) age ≥18 and (2) previously confirmed SARS-CoV-2 PCR in swab or sputum. Initial presentation of SARS-CoV-2-convalescent patients was scheduled 6 weeks after symptom onset or positive SARS-CoV-2 polymerase chain reaction testing (PCR), if asymptomatic. Each individual was invited for follow-up visits at month 4 and 7, regardless of symptoms and antibody status. At each visit, symptoms and serum SARS-CoV-2 immunoglobulins G were assessed. The study was approved by the Institutional Review Board of the UHC, Germany (16-054 and 20-1187).

### Long-term health consequences after mild infection were summarized as post-COVID syndrome (PCS)

At visits, individuals completed a medical history questionnaire and evaluated by a trained physician. For this purpose, systematic questionnaires for first (20 items) and follow-up (10 items) visits were developed. In addition, information gathered from the written questionnaires was supplemented with specific medical history and physical examination at each presentation. As most common symptoms, we summarized presence of long-term health consequences such as anosmia, ageusia, fatigue or shortness of breath at month 4 respectively 7 as post-COVID syndrome (PCS).

### SARS-CoV-2 IgG Serology

Euroimmun anti-SARS-CoV-2 ELISA for immunoglobulin class G targeting the S1 domain of the spike protein was used for serological testing (EuroimmunDiagnostik, Lübeck, Germany). According to manufacturer’s recommendations, IgG values were interpreted as positive if the ratio extinction value of patient sample and extinction value of the calibrator (S/CO-Ratio) was ≥1.1, borderline between <1.1 and ≥0.8 and negative <0.8.

### Statistical Methods

Clinical data and patient characteristics were systematically recorded in electronic case report forms (eCRF) using the online cohort platform ClinicalSurveys.net developed by UHC. Patient characteristics as absolute numbers and percentages or medians and interquartile ranges (IQR) are shown in Table 1. Uni-and multivariable logistic regression was used to identify predictors of PCS. Variables with p-values <.20 in the univariate regression were included in the multivariable model. Unadjusted and adjusted odds ratios (OR) with 95% confidence intervals (CI) from logistic regression were reported for various clinical data and patient characteristics at baseline. Multicollinearity issues were tested using the variance inflation factor (VIF). We used SPSS Statistics (IBM, Armonk, NY) and GraphPad Prism (GraphPad Software, La Jolla, CA) to compile analyses and graph the data.

**Table 1.**
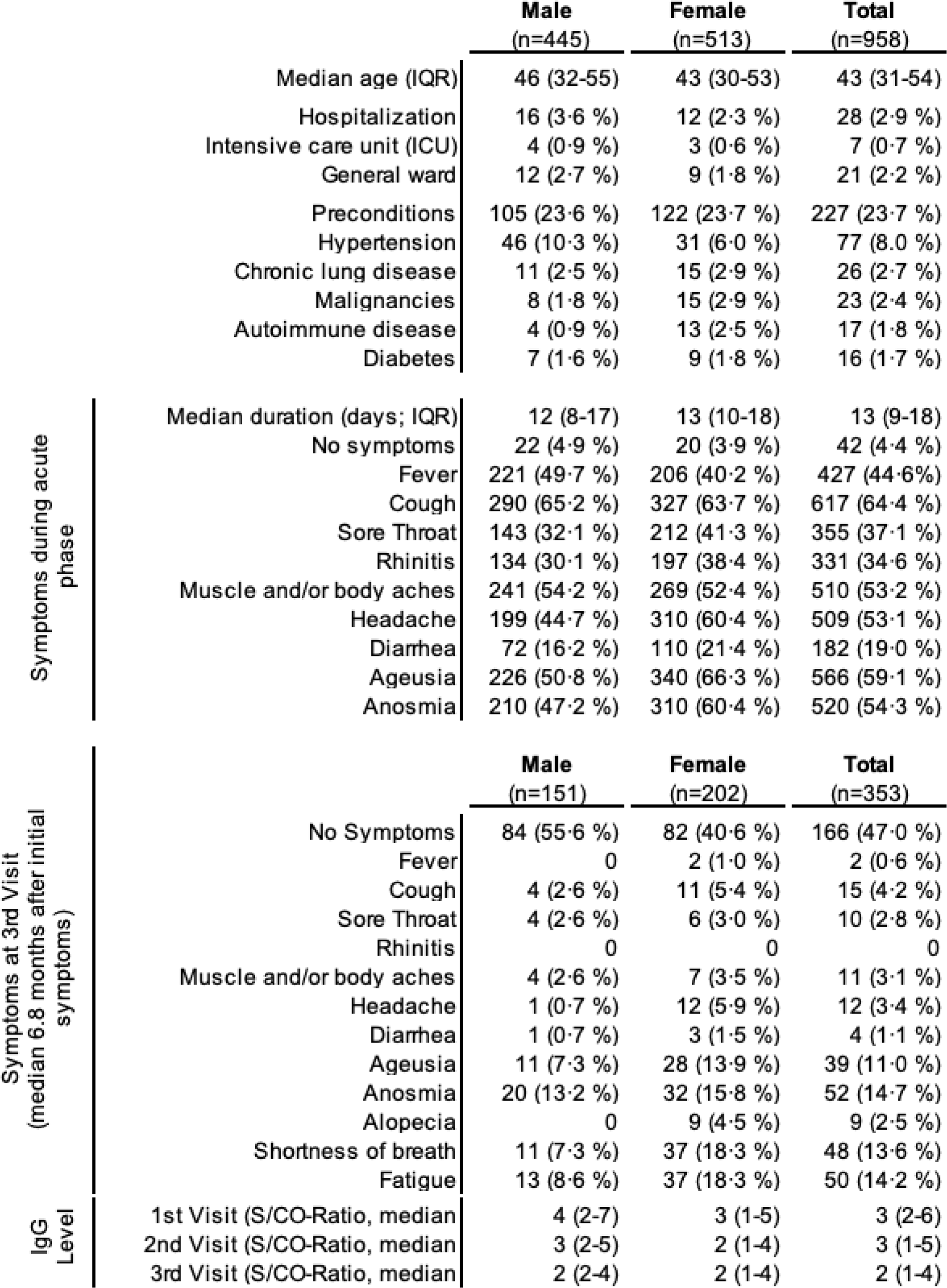
Patient characteristics of SARS-CoV-2 infected patients. Baseline and at third visit (T3, in median 6.8 months after initial symptoms (T0)). IQR, interquartile range; IgG, immunoglobulin class G.

### Results Participants

Between April 6^th^ and December 2^nd^ 2020, a total of 958 COVID-19-convalescent individuals presented to the post-COVID outpatient clinic of UHC, of which 442 were followed up until the second follow-up visit at 4.3 months (median 131 days (IQR 112-149) and 353 until the third follow-up visit at 6.8 months (median 207 days (IQR 187-234) (figure 1 and 2). According to WHO COVID-19 clinical progression scale ^4^, 97.1% of patients (930/958) initially presented with mild disease (score 1-3). Only 2.2% (21/958) and 0.7% (7/958) of all patients experienced moderate (score 4-5) or severe disease (score 6-9), respectively. The proportion of 46.5% men (445/958) and 53.5% women (513/958) was balanced (Table 1). Median age at the first visit was 43 years (31-54). Concomitant conditions prior to SARS-CoV-2 infection were reported in 23.7% of patients (227/958). Of these, the most frequent were hypertension 8.0% (77/958), chronic lung disease 2.7% (26/958) and malignancies 2.4% (23/958) (Table 1). Moreover, diabetes and autoimmune diseases were reported by 1.7% (16/958) and 1.8% of patients (17/958), respectively. The majority of patients was not hospitalized due to COVID-19 (97.1%, 930/958, Table 1). Only 2.9% (28/958) of all patients were hospitalized, of which 0.7% (7/958) were admitted to the intensive care unit and only 0.2% (2/958) were mechanically ventilated (Table 1). The remaining 2.2% (21/958) were admitted to the general ward (Table 1).

**Figure 2.**
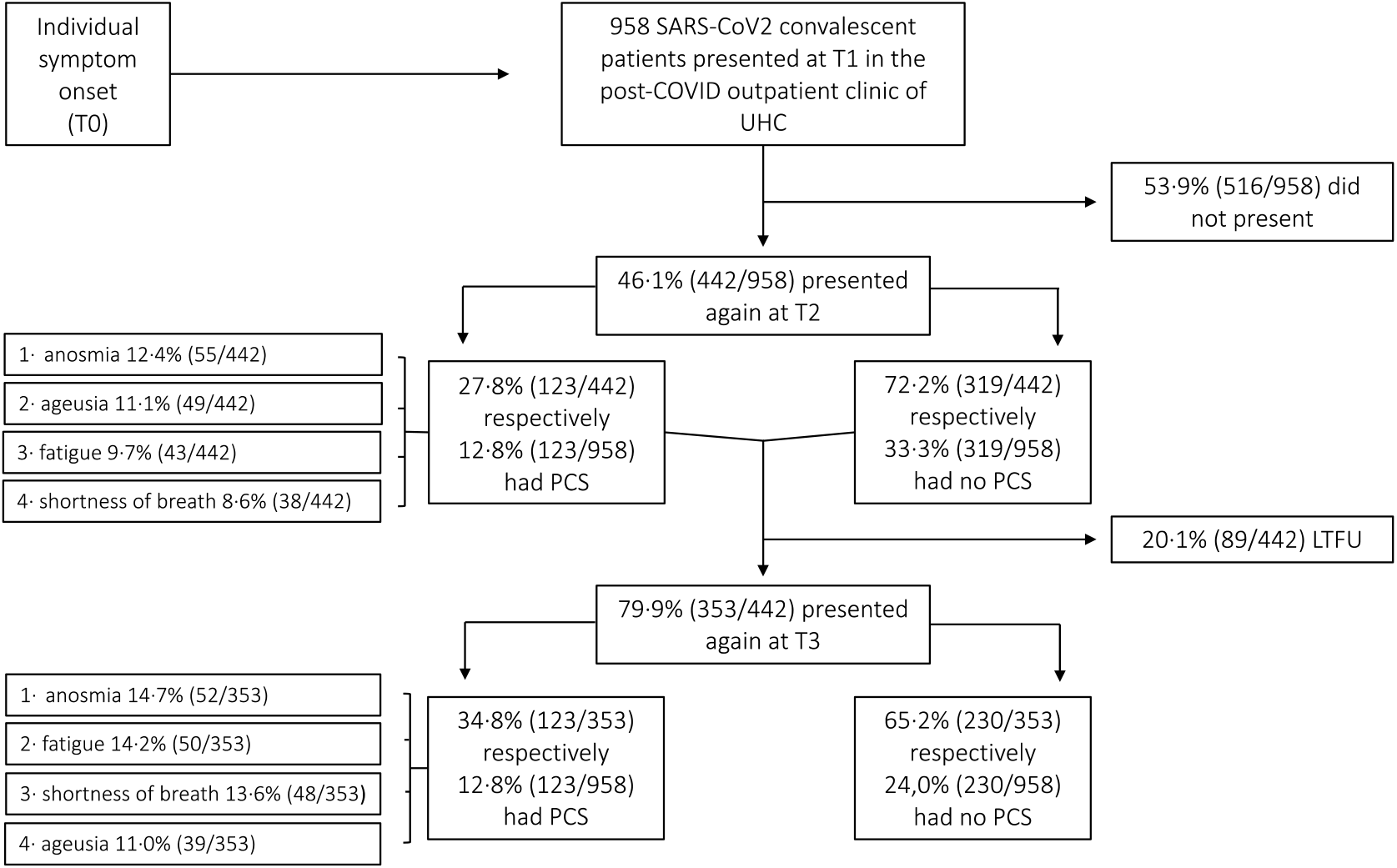
Distribution of post-COVID (PCS) syndrome among SARS-CoV-2 convalescent patients. UHC, University Hospital Cologne; T0, symptom onset; T1, first study visit; T2, second study visit; T3, third study visit; LTFU, lost to follow-up.

### SARS-CoV-2 serology

IgG titers (S/CO-Ratio) gradually decreased from first (T1) to third (T3) visit (median T1: 3 (IQR 3 – 4), T2: 3 (IQR 2-3) and T3: 2 (IQR 2-2). For further analyses, patients were stratified in three groups based on IgG titers at first visit: IgG low ≤1.1, IgG medium 1.2-4.0 and IgG high >4.0. Symptoms at third study visit were associated with low IgG titer at first study visit (figure 3).

**Figure 3.**
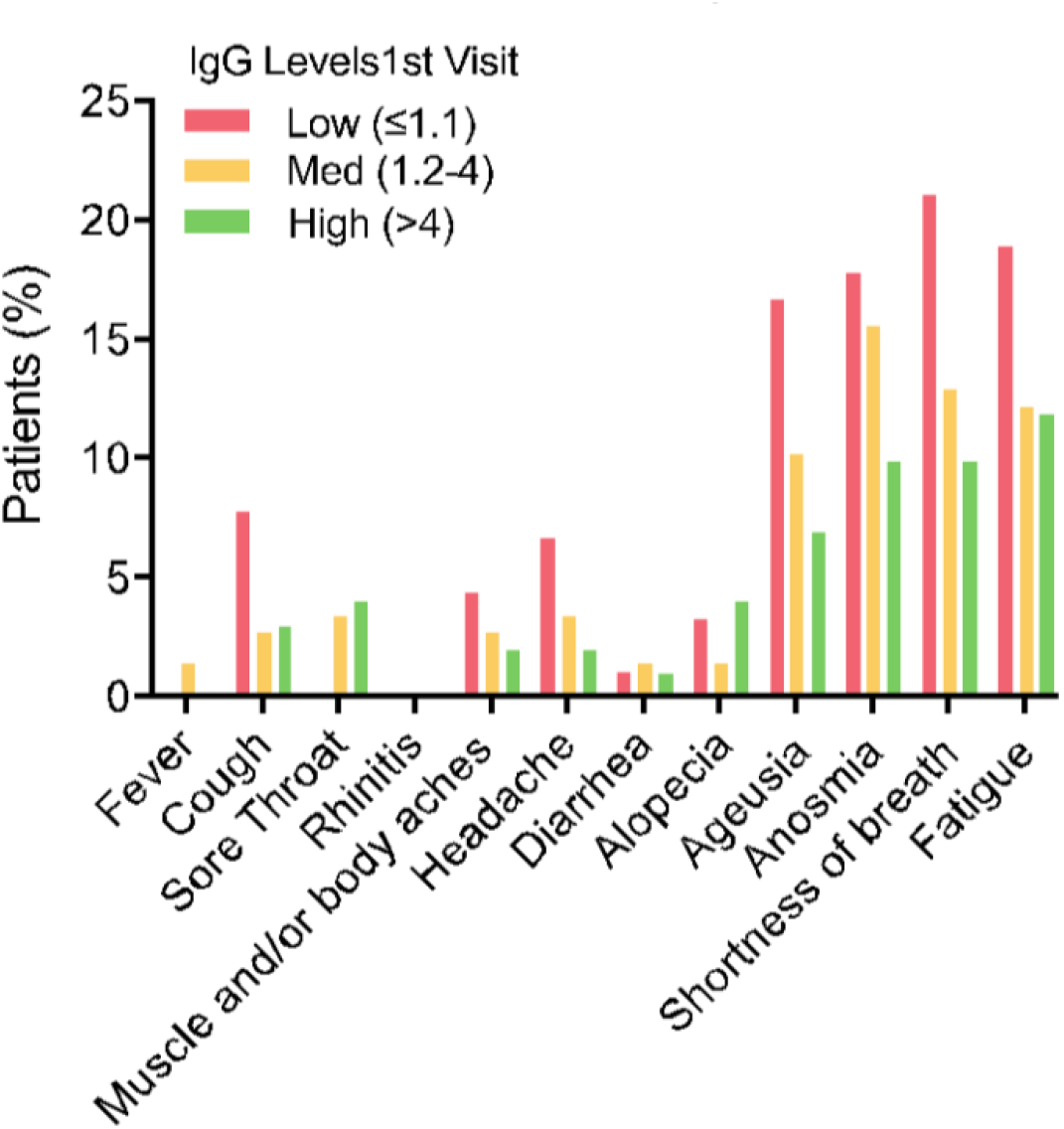
IgG levels at first visit are predictors for post-COVID syndrome (PCS) Bars show percentage of patients that reported the respective symptom at their third visit. Patients have been stratified by IgG levels at their first visit (IgG strata: low: ≤1.1, medium (med) 1.2-4, high >4, values are the ratio of extinction value of patient sample/extinction value of calibrator)

### Symptoms and predictors for PCS

Most common symptoms at disease onset were cough 64.4% (617/958), ageusia 59.1% (566/958), anosmia 54.3% (520/958), body aches 53.2% (510/958), headache 53.1% (509/958) and fever 44.6% (427/958) (Table 1). 4.5% (43/958) of SARS-CoV-2-infected patients were initially asymptomatic.

In contrast, most common symptoms after a median follow-up of 4 months (IQR 3-5) were 12.4% anosmia (55/442), 11.1% ageusia (49/442), 9.7% fatigue (43/442) and 8.6% shortness of breath (38/442) (figure 2). After a median follow-up of 7 months (IQR 6-8) symptoms still prevailed similarly : 14.7% anosmia (52/353), 13.6% shortness of breath (48/353), 14.7% fatigue (52/353) and 11.0% ageusia (39/353) (figure 2). Further symptoms present at 7 months were 3.7% headache (13/353), 2.5% alopecia (9/353) and 1.1% diarrhea (4/353).

The presence of any of these four symptoms (anosmia, ageusia, fatigue or shortness of breath) at month 4 respectively 7 was summarized as PCS. Using this approach, we identified PCS in 27.8% (123/442) of SARS-CoV-2-infected patients (Table 2) at month 4 and in 34..8% (123/353) of patients at month 7.

**Table 2.**
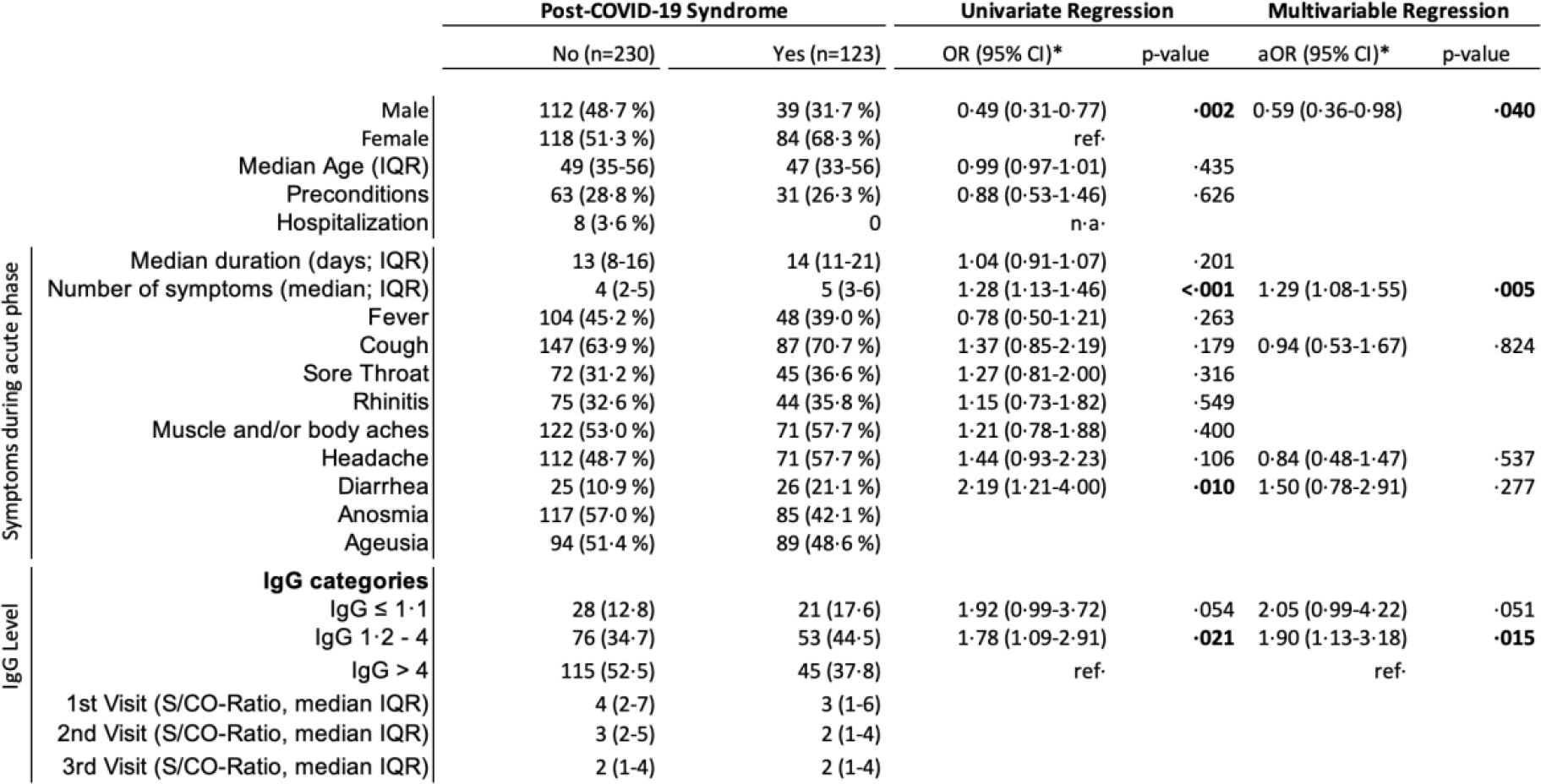
Predictors of post-COVID syndrome (PCS) based on an uni- and multivariable logistic regression model. *Odds-Ratio, 95% confidence interval and p-value obtained using binary logistic regression. n, number; OR, Odds-Ratio; aOR, adjusted Odds-Ratio; ref, reference; n/a, not applicable, IQR, interquartile range; S/CO-Ratio, signal-to-cutoff Ratio.

Univariate logistic regression revealed several factors and symptoms during acute COVID-19 that were associated with an increased risk of PCS after 7 months. Multiple symptoms (odds ratio (OR) 1.28; 95% confidence interval (95% CI): 1.13-1.46), diarrhea (OR 2.19; 95% CI 1.21-4.00), ageusia (OR 2.16; 95% CI 1.36-3.43), anosmia (OR 3.79; 95% CI 2.36-6.09) and baseline IgG titers between 1.2 and 4 (OR 2.06; 95% CI 1.19-3.53) were all predictors for a PCS after 7 months. Male gender was associated with a lower risk for PCS (OR 0.49; 95% CI 0.31-0.77). In the multivariable logistic regression model a lower baseline level of SARS-CoV-2 was associated with a higher risk of developing PCS after 7 months IgG (initial IgG 1.2-4; OR 2.06 (95% confidence interval (95%CI) 1.19-3.53), p=0.009 and initial IgG ≤1.1; aOR 2.05 (95%CI 0.96-4.37), p=0.054). Moreover, anosmia and diarrhea during acute COVID-19 were independent predictors for a PCS after 7 months with an aOR of 5.12 (95% CI 2.43-10.76, p=<0.001) and 2.35 (95%CI 1.13-4.90, p=0.023), respectively (Table 2).

## Discussion

In this large prospective study of 958 SARS-CoV-2-convalescent patients, the majority initially presented with absent to mild symptoms (WHO clinical progression scale score 1-3) ^4^. We observed that 27.8% (123/442) were found to have long-term health consequences after four months, defined by the presence of at least one symptom (anosmia, ageusia, fatigue or shortness of breath). In the total cohort this represented 12.8% (123/958). Health related events also persisted at month 7 in 34.8% patients (123/353), which represented 12.8% (123/958) of our total cohort. The most frequent concomitant conditions prior to SARS-CoV-2 infection were hypertension, chronic lung disease, and malignancies, and thus reflected the most common concomitant diseases in the general population^5-7^ as well as those reported within other studies with comparable SARS-CoV-2-convalescent cohorts ^8,9^. We have complete clinical and serological data of 353 patients out of initially 958 patients after 7 months. High drop-out rate of 20% between 4 and 7 months suggests that rate of long-term symptoms may, in fact, be lower than 27.8% (123/442) and 34.8% (123/353) at 4 and 7 months, respectively, assuming those patients lost to follow-up were no longer symptomatic. Hence, considering our total cohort of 958 patients, we can therefore conclude that 12.8-27.8% of patients suffer from long-lasting symptoms. To the best of our knowledge this is the first set of data, in which a large cohort of SARS-CoV-2-convalescent patients was prospectively followed up with a standardized procedure questionnaire for a median duration of over 7 months.

Lately, it has become evident that some patients continue to experience symptoms weeks and months after onset of COVID-19, regardless of disease severity ^2,10-12^. To date, neither this new disease entity itself nor the exact terminology has been defined. So far, the terminology has ranged from “long COVID”^11^, “chronic COVID syndrome”^13^ to “post-COVID syndrome”^10^ or “post-acute COVID-19 syndrome”^14^. Since the symptoms persist even after 7 months, we decided to use the terminus post-COVID syndrome (PCS) rather than post-acute COVID-19 syndrome.

Moreover, a clinical case definition of PCS has not been clearly specified ^13^. In particular, most data was derived from hospitalized patients with severe disease ^11,15,16^. In our study population 97.1% (930/958) were not hospitalized during acute phase of COVID-19, which is consistent with hospitalization rate in our region ^3^. In recent research, time span to define the syndrome varied between >28-30 days ^12,17^, 60 days ^12,16,17^ or longer than 3 months ^12,18^. However, time frames were based on maximum follow-up time of the respective cohorts rather than critical evaluation of actual symptoms. Previously, it has also not been systematically evaluated which symptoms characterize PCS. Shortness of breath ^16^, fatigue ^16,19,20^, joint pain ^16^, anosmia and ageusia ^2^ are symptoms most commonly reported and therefore associated with PCS. Worsened quality of life was observed in 2/3 of hospitalized patients^2,16^.

Most studies focusing on long-term symptoms included hospitalized COVID-19 patients that often continued to experience symptoms for 1-2 ^2,16^ or 6 months ^15^ after disease onset. Unlike patients with mild COVID-19, hospitalized patients are more prone to long-term symptoms, which are a result of mechanical ventilation or prolonged immobilization, but are not COVID-19 specific. These patients are consequently expected to have longer convalescence. Sudre et al. reported on 4182 COVID-19 patients, who entered their symptoms in a digital diary ^12^. In this cohort, symptoms were reported for more than 28 days in 13.3% of patients, and for more than 12 weeks in 2.3%. Huang et al recently reported that 6 months after being hospitalized with COVID-19, 63% of a total of 1655 patients suffered from fatigue or muscle weakness and 26% experienced sleep difficulties ^9^.

In contrast to previous reports, our study has several key advantages: i) Our cohort consists of mostly mild COVID-19 cases that have been prospectively followed for a median time of 6.8 months. ii) All patients were seen by at least one trained physician at each visit to critically assess all reported symptoms and to supplement information gathered from the written questionnaires by specific medical history and physical examination. iii) SARS-CoV-2 IgG were assessed at each visit allowing to correlate reported symptoms with serological data.

Using this approach, we identified fatigue, anosmia, ageusia and shortness of breath as most common symptoms in PCS patients. Fatigue has previously been reported as one of the most common symptoms of PCS^18,19^ and among survivors of SARS-CoV-1 pandemic in 2003^21^. A possible physical correlate for fatigue could be the endothelial dysfunction in brain capillaries recently described in Nauen et al.^22^. In line with data published by Townsend et al., we observed that even patients with initially mild disease may develop fatigue as a leading symptom of PCS^18^. It should be noted, however, that fatigue can vary from person to person and that no single test confirms a diagnosis of fatigue. Female patients and individuals with a prior diagnosis of depression or anxiety had a higher risk of suffering from fatigue^18^.

Post-infectious syndrome, especially paired with fatigue, has already been associated with different pathogens such as Epstein-Barr virus, Coxiella burnetii or Borrelia burgdorferi often lasting over 6 months ^23,24^. Fatigue may also be related to autoimmune diseases, for instance sarcoidosis ^25^. In the study by Sudre et al. ^12^, long-term complaints were characterized by symptoms of headache, fatigue, dyspnea and anosmia. Furthermore, occurrence of a PCS was associated with older age, higher body mass index and female gender. In contrast, patients with severe and acute COVID-19 showed disproportionate frequency of being male, nonwhite, and were more likely to be obese ^26,27^. In line with previously published data, our results indicate that women are more likely to be affected by PCS. However, this may be because women are more prone to report their symptoms to a doctor^28^.

In addition, anosmia is a well described symptom in COVID-19 patients ^9,29,30^ that appears to be more common in women ^8,31^ and lessens over time^8^. The pathophysiological mechanisms of anosmia are not yet fully understood. As SARS-CoV-2 enters the nasal epithelium via the Angiotensin-converting enzyme 2 (ACE-2) receptor^32^, injuries in the olfactory neuroepithelium are possible.

Finally, in contrast to other viral infections with respiratory pathogens such as the common flu, observed shortness of breath and anosmia in the context of PCS may possibly be due to pathomechanism of SARS-CoV-2: vascular angiogenesis was observed at a greater frequency in lungs from patients with COVID-19 when compared to patients infected with influenza virus_33_.

Besides, we also observed that diarrhea during acute COVID-19 was associated with a higher risk of developing PCS. Gaebler et al recently examined intestinal biopsies obtained from asymptomatic individuals 3 months after COVID-19 onset. They showed persistence of SARS-CoV-2 in the small intestine of 7 out of 14 subjects ^34^. Since the intestine is the largest lymphatic organ^35^, there may be a possible link between residual viral particles in the intestine and long-term symptoms.

In addition to clinical evaluation, SARS CoV-2 IgG titers were determined on a regular basis. A lower SARS-CoV-2 IgG titer at the beginning of the observation period was associated with a higher frequency of PCS. This trend was also identified in individuals with IgG lower than 1.1. Overall, low IgG titers six weeks after symptom onset could result in an insufficient humoral immune response, possibly leading to PCS in the long term in these patients.

The correlation between the level of IgG titers and disease severity during the acute phase of COVID-19 is well described, as patients with a severe course of COVID-19 show higher levels of anti-spike 1 (S1) IgA and IgG than patients with mild course^36-38^. Disease severity also correlates with more potent SARS-CoV-2 neutralizing antibody response. Moreover, in cases with mild disease, older age, higher IgG levels and fever are associated with the development of SARS-CoV-2 neutralizing activity (Vanshylla et al., submitted). IgG antibody levels have been shown to decay over the first weeks but are still detectable in the majority of patients (87%) 10 months after the individuals have recovered from mild COVID-19 (Vanshylla et al., submitted). In contrast, no data are available on possible association between a low IgG titer and a PCS.

Our study has limitations, as common laboratory parameters such as c-reactive protein (CRP), D-Dimers, creatinine, etc. were not routinely assessed. A high drop-out rate within 7 months suggests that true rate of PCS may be lower than 34.8%, as asymptomatic patients may not show up for later follow-up visits. Besides, smoking was not specifically queried, a factor that is known to be associated with poorer outcomes of acute COVID-19 ^39^. Symptoms like fatigue or shortness of breath have initially neither been assessed by validated scoring methods nor objectivated by examinations, since at that time, the rapid acquisition of reconvalescent plasma donors was top priority. However, both of these symptoms were critically discussed between the patient and physician at each clinic visit using the respective completed systematic questionnaire. The symptoms anosmia and ageusia were solely investigated by questionnaires and not by validated olfactory or gustatory test. Whether these patients really had a “loss” or rather a reduction (hyposmia/hypogeusia) or change (e.g. parosmia) of these senses, thus remains unclear. With persistent symptoms even four months after initial onset of symptoms, the focus of the outpatient clinic was shifted and, if necessary, further examinations such as lung spirometry, spiroergometry, olfactometry and laboratory diagnostic examinations were carried out, which are not included in this report. Failure to assess sociodemographic, clinical and laboratory factors as potential confounders may have biased the results of our multivariable model and led to inaccurate conclusions.

Taken together, we propose that PCS is characterized by fatigue, anosmia, ageusia or shortness of breath, which cannot be explained by an alternative diagnosis. Symptoms persist after 4 to 7 months post-infection. These symptoms are more frequent in SARS-CoV-2 infected female patients and are associated with lower serum IgG titers, anosmia and diarrhea at disease onset. It should be emphasized that PCS can occur even after a very mild initial phase of COVID-19 in outpatient setting. As up to 81% of all SARS-CoV-2 infected patients present with mild disease ^40^, it can be expected that PCS will affect a larger number of individuals than initially assumed, posing major medical, social and economic challenges. Even following mild courses of SARS-CoV-2 infection, 11% of patients in our cohort still could not fully participate in everyday and work life 7 months after disease onset, which highlights the potential economic damage of PCS.

The full clinical picture of PCS is complex and far from being understood. However, the viral tropism defined by the entry into cells through a widely expressed ACE2 receptor^32^, makes it highly possible that many organs have the potential to undergo not only acute, but also chronic damage, adding to the very diverse clinical picture of PCS^41^. Efficient or causal therapies have not yet been identified. In our own experience, specific and general methods of physical rehabilitation can be helpful. We conclude the establishment of specialized interdisciplinary post-COVID outpatient clinics is needed to provide individualized care, as well as conduct further research and develop new therapeutic options for patients with post-COVID syndrome.

## Data Availability

All data referred to in the manuscript are available at all time.

## Author contributions

MA, PS, MS, IS and CL developed the first draft outline and wrote parts of the manuscript. Subsequent drafts were developed by MH, FK, HG, LG, FD, CH, VP, KV, TR, MR, LO, NT, SR, CA, JCL, BG, VDC and GF. PS, MS, MA, IS, KV, VDC and CL analyzed and collected data, and interpreted the findings. All authors were involved in either patient care or development of post-COVID outpatient clinic of UHC. All authors contributed to all sections relevant to their experience and helped finalize the text and content. MA, PS, IS and CL contributed equally to this manuscript.

